# Single-Cell RNA Sequencing Reveals the Altered Landscape of Immune Cells in Immune Checkpoint Inhibitor Related Myocarditis

**DOI:** 10.1101/2022.01.21.22269639

**Authors:** Bowen Lou, Manyun Guo, Fangyuan Chen, Chen Wang, Gulinigaer Tuerhongjiang, Tao Zheng, Bo Zhou, Zuyi Yuan, Jianqing She

## Abstract

**Background:** Myocarditis has emerged as a rare but lethal Immune checkpoint inhibitor (ICI)-associated toxicity. However, the exact mechanism for ICI related myocarditis remains underexplored; and the specific therapeutic targets is still lacking. In this study, we used scRNA-seq to characterize the transcriptomic profiles of single cells from the peripheral blood mononuclear cell (PBMC) of ICI related myocarditis during fulminant myocarditis and disease recovery.

**Methods:** PBMC samples were taken from the patient during fulminant ICI related myocarditis and after disease remission. Cells were isolated from blood samples by density gradient centrifugation over Ficoll-Paque. Single-cell RNA sequencing with 10X genomics was performed. Subpopulation determination, functional analysis, single-cell trajectory and cell-cell interaction analysis were carried out afterwards.

**Results:** We presented the altered landscape of immune cells and differential genes in ICI related myocarditis during the disease activity and remission using scRNA-seq. Substantial immune cell composition and intercellular communication were found to be altered. Monocyte, NK cell as well as B cell subpopulations contributed to the regulation of innate immunity and inflammation in ICI related myocarditis. T cell subpopulations highly expressed genes associated with PD-1 inhibitor resistance and hyper-progressor. At last, the intercellular communication in ICI related myocarditis was significantly dysregulated.

**Conclusion:** By identifying altered pathways and highlighting a catalog of marker genes, this study has revealed the diversity of cellular populations in ICI related myocarditis, marked by their distinct transcriptional profiles and biological functions. Our investigation would shed light on the pathophysiological mechanism and potential therapeutic targets of ICI related myocarditis in continuous exploration.

## Introduction

With the revolutionized utilization of Immune checkpoint inhibitor (ICI) therapy in various malignancies, myocarditis has emerged as a rare but lethal ICI-associated toxicity, the mortality rate of which approaching 50%. While stimulating an effective tumor response by blocking the co-inhibitory pathway used by tumor cells to evade the T cell-mediated immune response, ICI could also elicit profound immune reaction and cytokine release, causing significant systemic abnormalities, including fulminant myocarditis^1, 2^. However, the exact mechanism for ICI related myocarditis remains underexplored; and the specific therapeutic targets is still lacking. As ICI functions mainly by interfering T cell-mediated immune response, to investigate an atlas of peripheral blood cells and characterize their respective functions are essential for understanding the pathophysiology and developing treatment strategies for ICI related myocarditis.

ScRNA-seq has emerged as a powerful tool for defining cellular components and functions in various diseases and tissues across a spectrum of tumor and cardiovascular disease. Recent studies have investigated comprehensive single-cell landscape of immune cells among Programmed Death Molecule 1 (PD-1) treated patients as well as animal models. It has been revealed that the hepatitis induced by combination of ICIs is associated with a robust immune activation signature in all subtypes of T cells; together with a central role for liver monocyte-derived macrophages in promoting a pro-inflammatory T cell response^3^. ScRNA-seq has also revealed that the homing macrophages expressing unique scavenger molecules are required for the effect of Programmed Cell Death 1 Ligand 1 (PD-L1) blockead^4^. Moreover, scRNA-seq has further links the durable responses to ICI with T cell receptor (TCR) repertoire^5-7^.On the other hand, a comparison of scRNA-seq data from different autoimmune myocarditis models suggests that some cell clusters such as macrophage cluster and T-helper cells are associated with the inflammatory response in the myocarditis^8^. However, data on the single-cell transcriptional profiles and immunological network of ICI related myocarditis during the disease onset and remission remain unknown.

In this study, we used scRNA-seq to characterize the transcriptomic profiles of single cells from the peripheral blood mononuclear cell (PBMC) of ICI related myocarditis during fulminant myocarditis and disease recovery. Our results revealed the diversity of cellular populations in ICI related myocarditis, marked by their distinct transcriptional profiles and biological functions. Furthermore, we described the similarities and differences in the cellular composition during myocarditis onset and recovery. Our study could serve as an important resource for studying the potential mechanism and pathophysiology of ICI related myocarditis.

## Methods

### Study design and eligibility

This study was designed to profile transcriptomic changes in PBMC samples during fulminant ICI related myocarditis and during disease remission. PBMC samples were taken from the patient with myocarditis before any treatment and 28 days later when the cardiac lesion markers, echocardiography and inflammatory markers returned to normal (Supplementary Fig 1). The ICI related myocarditis happened 21 days after the utilization of Tislelizumab, a PD-1 inhibitor as a treatment for lung cancer. The diagnose of ICI related myocarditis was based on clinical symptoms, echocardiography and laboratory markers according to the current guidelines^9, 10^. Written informed consent was obtained from the patient, with ethnic committee approval at the First Affiliated Hospital of Xi’an Jiaotong University.

### PBMC collection and library preparation for single-cell RNA sequencing

PBMCs were isolated from blood samples by density gradient centrifugation over Ficoll-Paque, and cryopreserved in 10% dimethyl sulfoxide in fetal bovine serum and stored in liquid nitrogen. Single-cell suspensions were loaded to 10x Chromium to capture 5000 single cells according to the manufacturer’s instructions of 10X Genomics Chromium Single-Cell 3’kit (V3). The following cDNA amplification and library construction steps were performed according to the standard protocol. Libraries were sequenced on an Illumina NovaSeq 6000 sequencing system (paired-end multiplexing run,150bp).

### Single-cell RNA sequencing

Quality control and data processing were carried out by LC-Bio Technology (Hangzhou, China). 10X genomics raw data were processed by using the Cell Ranger Single-Cell Software Suite (release 5.0.1), including using cellranger mkfastq for demultiplexing raw base call files into FASTQ files and then using cellranger count for alignment, filtering, barcode counting, and UMI (unique molecular identifier) counting. The reads were aligned to the Homo_sapiens_GRCh38_96 reference genome by using a prebuilt annotation package downloaded from the 10X Genomics website. The outputs from different lanes were finally aggregated by using cellranger aggr with the default parameter settings.

We mapped UMIs to genes, followed by removing low-quality cells. Cells were flagged as poor-quality if 1) the number of expressed genes was < 500 and 2) 25% or more UMIs mapped to mitochondrial or ribosomal genes. Cells meeting the latter threshold were usually nonviable or apoptotic. As a result, 8183 and 9546 cells were obtained from fulminant myocarditis and recovered myocardial functions status, respectively. The data were then normalized. Genes with high variation were identified using Seurat function FindVariableFeatures.

Principal component analysis(PCA) for dimensional reduction was performed using Seurat function RunPCA. We then used the FindNeighbors and FindClusters (resolution = 0.8)functions in the Seurat package for cell clustering analysis and displayed the 2D map using tSNE.

### Subpopulation Analysis

The FindAllMarkers function in Seurat was used to identify unique cluster-specific marker genes. We first identified PBMC clusters with the dominant expression pattern of cell markers, including monocyte, NK cell, CD4+ T cell, CT8+ T cell, B cell and megakaryocyte, from the online database and published literature. Putative clusters with common canonical markers were integrated into one cluster. Violin plots, heatmaps, and cluster annotations were carried out in Seurat with the corresponding footnotes.

### Functional Analysis

To identify the potential biological functions of cell clusters, we performed enrichment analysis with marker genes for cell clusters. Marker genes detected in >10% cells in a cluster with average log2FC>=0.26,P<=0.01 were used in the analysis. Enrichment analyses were performed using the OmicStudio tools at https://www.omicstudio.cn/tool. Cell subpopulations were annotated based on their expressed marker genes, functions, and proportions.

### Single-cell trajectory analysis

Single-cell trajectories were constructed by using the Monocle package based on the analysis of the UMI read data. The default settings were used for all other parameters. Pseudotime analysis data were classified into microglial clusters and cell states in different groups.

### Cell-cell interaction

We predicted intercellular communication network among different cell populations using CellPhoneDB(https://pypi.org/project/CellPhoneDB/). In brief, it uses the UMI count matrix to predict potential interactions of ligand and receptor between each pair of cell populations. The nodes of the network are ligand, receptor, and ECM molecules. The edges of the network are interaction between two molecules. Higher expression levels of ligands and receptors result in larger scores, and imply higher probability and stronger strength of interreactions. We used marker genes with log-fold-change >0.5 in each cell populations to refine the network. The results show the potential specific cell-cell interreactiontion pathways between two cell populations. To visualize the communication, top 20 ligand-receptor pairs based on predicted interaction scores were selected and plotted.

### Statistical Analysis

All statistical analysis was performed in R (Version 3.5.2). P<0.01 was considered as statistical significance. The significance of PCs in the dataset was determined using Seurat function JackStraw via a permutation approach. Differential proportion analysis of cell populations or subpopulations was performed by logit transformation of cell ratios in PBMC, and then t test between each pair. Differential expression of genes between cell clusters were determined with the Wilcoxon rank-sum test using Seurat FindAllMarkers. Overrepresentation tests were performed using clusterProfiler function enrichGO, and P were computed with hypergeometric tests. For differential proportion, differential expression, and enrichment analyses, P were adjusted for multiple hypothesis testing using the Benjamini-Hochberg method.

### Availability of Data

The data that support the findings of this study including scRNA-seq data and R scripts for single cell data analysis are available from the corresponding author upon reasonable request. Besides, the RNA-Seq datasets produced in this study are uploading to GEO (Gene Expression Omnibus, NIH) and the accession number will be accessed soon.

## Results

### ScRNA-seq atlas of cell populations in ICI related myocarditis and recovery phase after PD-1 initiation

To explore the profiling of specific cell populations at the single-cell level, peripheral blood mononuclear cell samples were collected during fulminant ICI related myocarditis onset and after disease remission. A total of 17729 cells were included in the subsequent analysis after quality filtering (Fig 1, A). We identified 17 cell clusters including monocytes (4 cell clusters), neutrophils (3 cell clusters), T cells (5 cell clusters), natural killer (NK) cells (6 cell cluster), B cells (1 cell clusters), and platelets (1 cell cluster) using unsupervised graph-based clustering and SingleR (Fig 1, B, Supplementary Fig 2). Each cell cluster exhibited differential percentage (Fig 1,D), indicating that the cellular components of the immune system are fundamentally altered during ICI myocarditis and recovery phase. Different cell types were characterized with marker genes and specific gene expression patterns, such as CD14 (Monocyte), NCAM1(NK Cell), CD4 (CD4+ T cell), CD8A (CD8+ T cell) and CD19 (B Cell) (Fig 1, C and Fig 2, A). The AREG, NRGN, CXCR4, IL1R2, and RBM38 were the top 5 significantly down-regulated genes, and CD8B, S100A12, MNDA, S100A9, TYMP mostly upregulated genes during myocarditis. The violin plots of the relative levels and the location in different cell clusters were shown in Fig 2, B. The NRGN, IL1R2, S100A12, MNDA, S100A9 and TYMP were generally expressed in different clusters of NK cells; CXCR4, RBM38 in NK, T and B cells, AREG in NK cells and CD8B in T cells.

**Figure 1.**
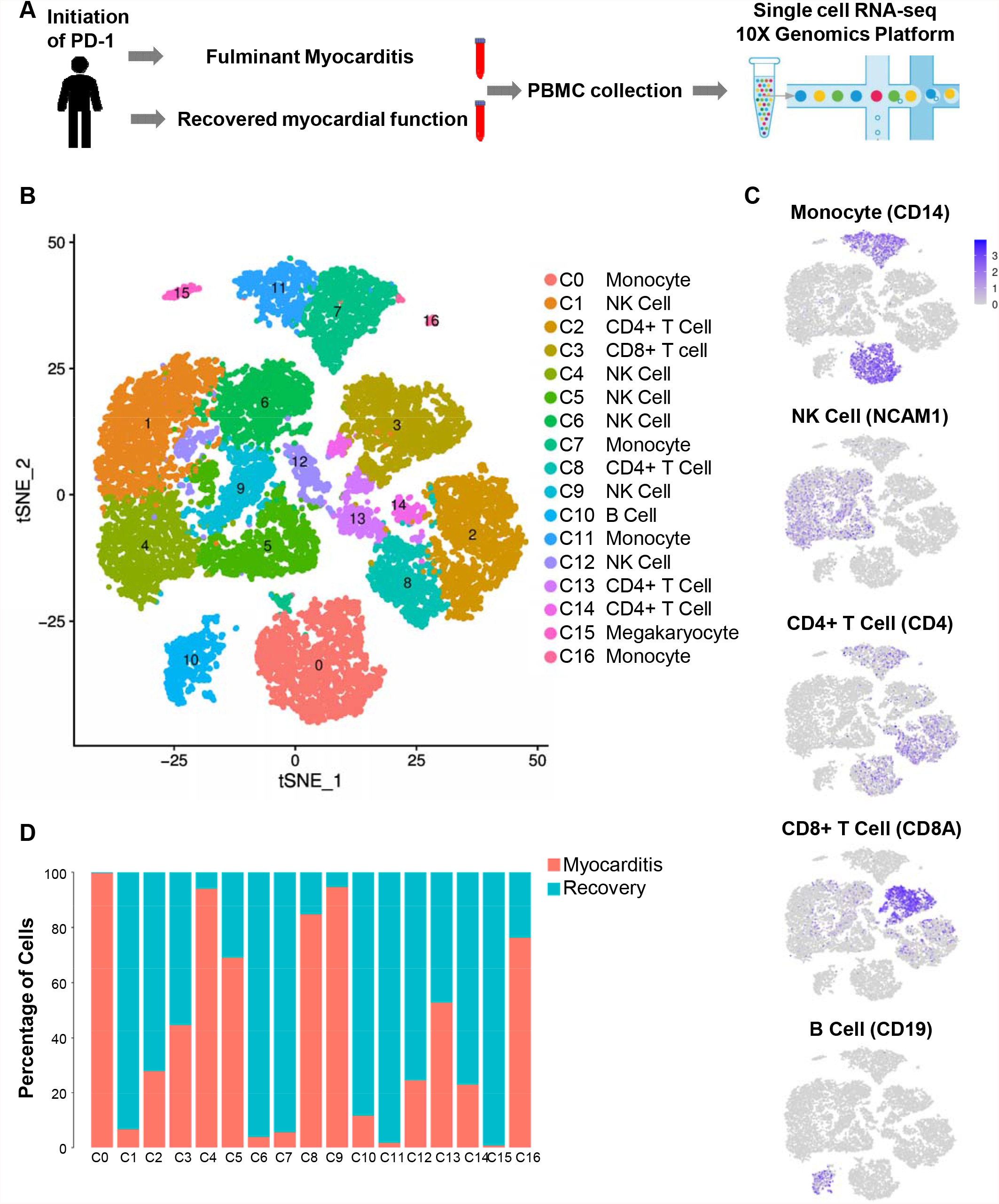
scRNA-seq atlas of cell populations in ICI related myocarditis and recovery phase after PD-1 initiation. A. Schematic diagram of PBMC collection during fulminant ICI related myocarditis onset and after disease remission; B. tSNE visualization of individual cell clusters from PBMC of ICI myocarditis and recovery phase using unsupervised graph-based clustering and SingleR; C. tSNE plots highlighting the marker gene of each cluster and cell-type; D. Differential percentage of each cluster between ICI related myocarditis and recovery phase.

**Figure 2.**
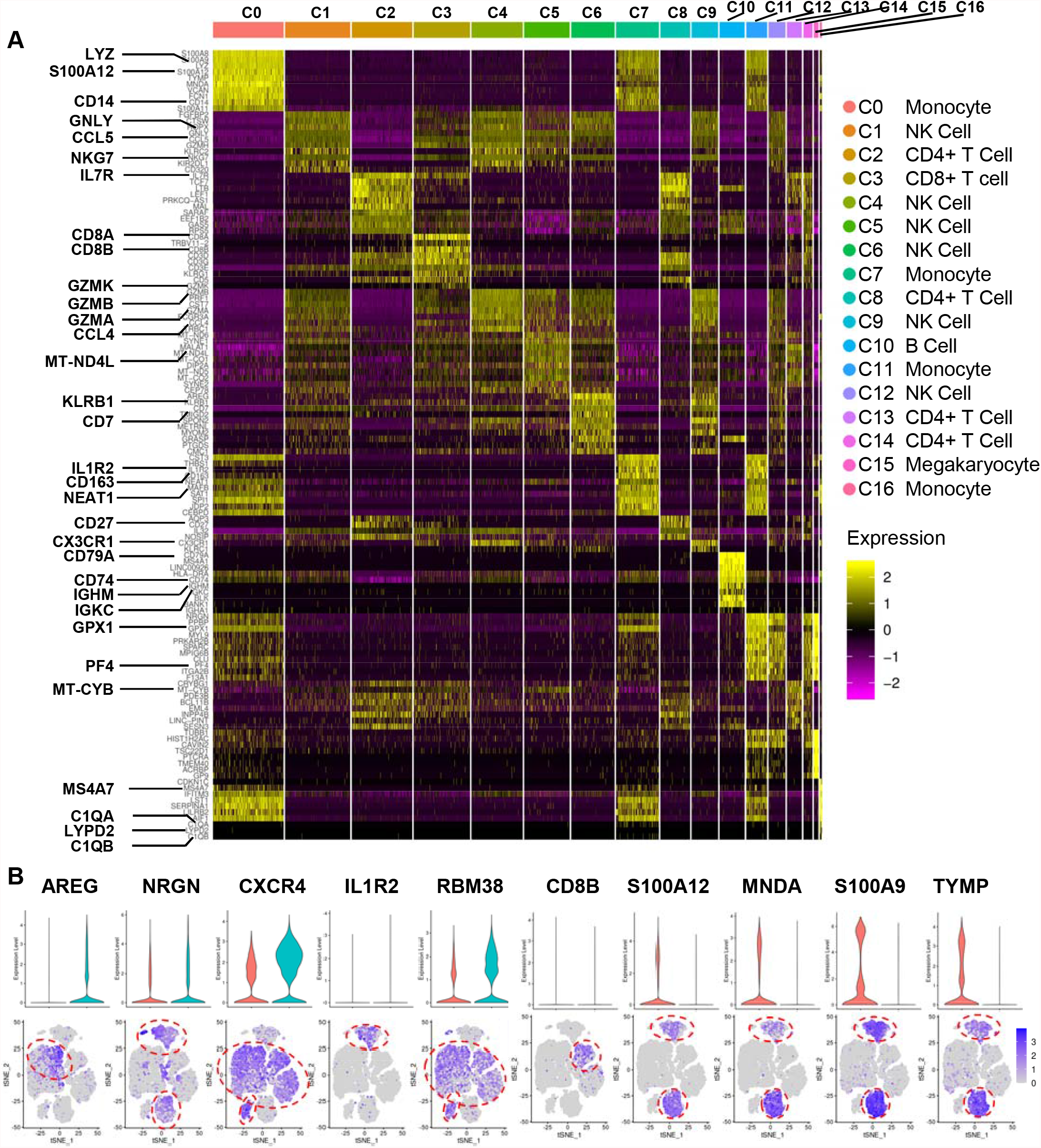
Top 10 markergenes and top 5 upregulated and downregulated genes in ICI related myocarditis and recovery phase after PD-1 initiation. A. Heatmap of Top 10 differentially expressed genes among all detected subclusters; B. The violin plots of top 5 upregulated and downregulated genes in ICI related myocarditis(right, red column) and recovery phase(left, blue column) after PD-1 initiation in different cell clusters.

### Monocyte subpopulations play inflammatory roles during ICI related myocarditis

We detected 4076 monocytes, which were further spilt into 4 clusters (Fig 1, A). The cluster C0 (Mono_1, S100A8^+^), C7 (Mono_2, NEAT1^+^), C11 (Mono_3, NRGN^+^) and C16 (Mono_4, LST1^+^) made up 52.0%, 31.1%, 15.4%, and 2.5% of the entire monocyte population. The clusters were identified to have substantially different proportion during the ICI myocarditis and recovery phase (Fig 3, A), suggesting a distinct transcriptomic profile. The Mono_1 (S100A8^+^) were predominant in ICI related myocarditis, while Mono_2 (NEAT1^+^) and Mono_3 (NRGN^+^) predominant in recovery phase (Fig 3, A). The gene expression pattern was different among monocyte subpopulations, and the marker genes of each subpopulation were shown in Fig 3, B.

**Figure 3.**
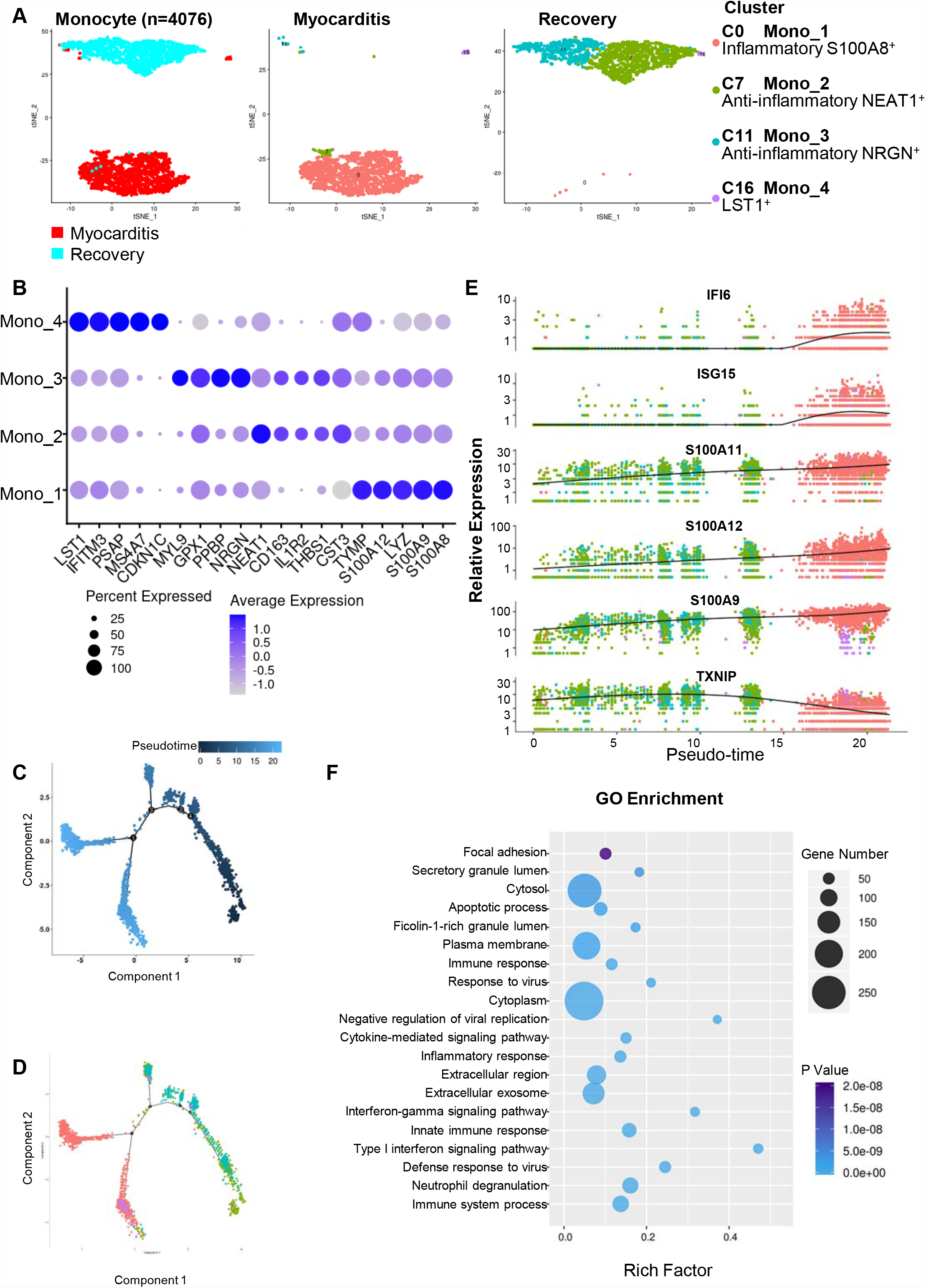
Monocyte subpopulations and its function in ICI related myocarditis and recovery phase. A. tSNE plots of different monocyte subpopulations during the ICI myocarditis and recovery phase, each color corresponds to different monocyte subpopulation/subtype as the right panel showed; B. Dot plot showing the markers of each monocyte subpopulation. Dot size corresponds to proportion of cells within the group expressing each gene, and dot color corresponds to expression level; C.Trajectory analysis of monocytes using Mononcle2. Dot color corresponds to pseudotime; D. Pseudotime trajectory analysis of four subtypes of macrophages by using Mono_2 (NEAT1^+^) as the root, each point indicates one single cell; E. Expressions of differential marker genes across four subtypes of monocytes; F. GO term enrichment analysis of main monocyte functions during ICI myocarditis.

Trajectory analysis using Mononcle2^11^ arranged most of monocytes into a major trajectory (Figure 3, C). Using Mono_2 (NEAT1^+^) as the root of the trajectory, we found that 2 cellular biology process decision points were dominated by the anti-inflammatory Mono_3 (NRGN^+^), and inflammatory Mono_1 (S100A8^+^) in pseudotime (Figure 3, D). This indicated that upon stimulation of PD-1, monocyte cells undergone pseudotime changes to either inflammatory or anti-inflammatory endpoints. Moreover, based on the differentiation time indicated by the pseudotime trajectory, the gene expression distribution map with 6 predominately differential genes were shown in Figure 3, E. Gradually significantly increased genes during pseudotime trajectory included IFI6, ISG15, S100A11, S100A12, and S100A9; and decreased gene TXNIP.

Additionally, the differentially expressed genes between ICI myocarditis and recovery phase were used for GO term enrichment analysis to predict the potential biological process that monocyte took part in during ICI myocarditis. Overall, the monocyte functions were mainly enriched in immune response, cytokine signaling and neutrophil degranulation, etc. during ICI myocarditis (Fig 3, F).

### NK cell subpopulations and different clusters contribute to the regulation of innate immunity in ICI related myocarditis

NK cell subpopulations separated into 6 subpopulations by first-level t-distributed stochastic neighbor embedding analysis (Fig 4, A). C4 (NK_2, highly expressing CST7, GZMA and GZMB), as well as C5 (NK_3, highly expressing MALAT1, MT-CO1 and MT-ND4L) were predominant in ICIs myocarditis, accounting for 39.04% of the entire NK cells. C1 (NK_1, highly expressing CCL5, and FGFBP2), as well as C6 (NK_4, highly expressing AREG and CD7) composed majority of NK cells during recovery, accounting for 43.88% of the entire NK cells (Figure 4, B).

**Figure 4.**
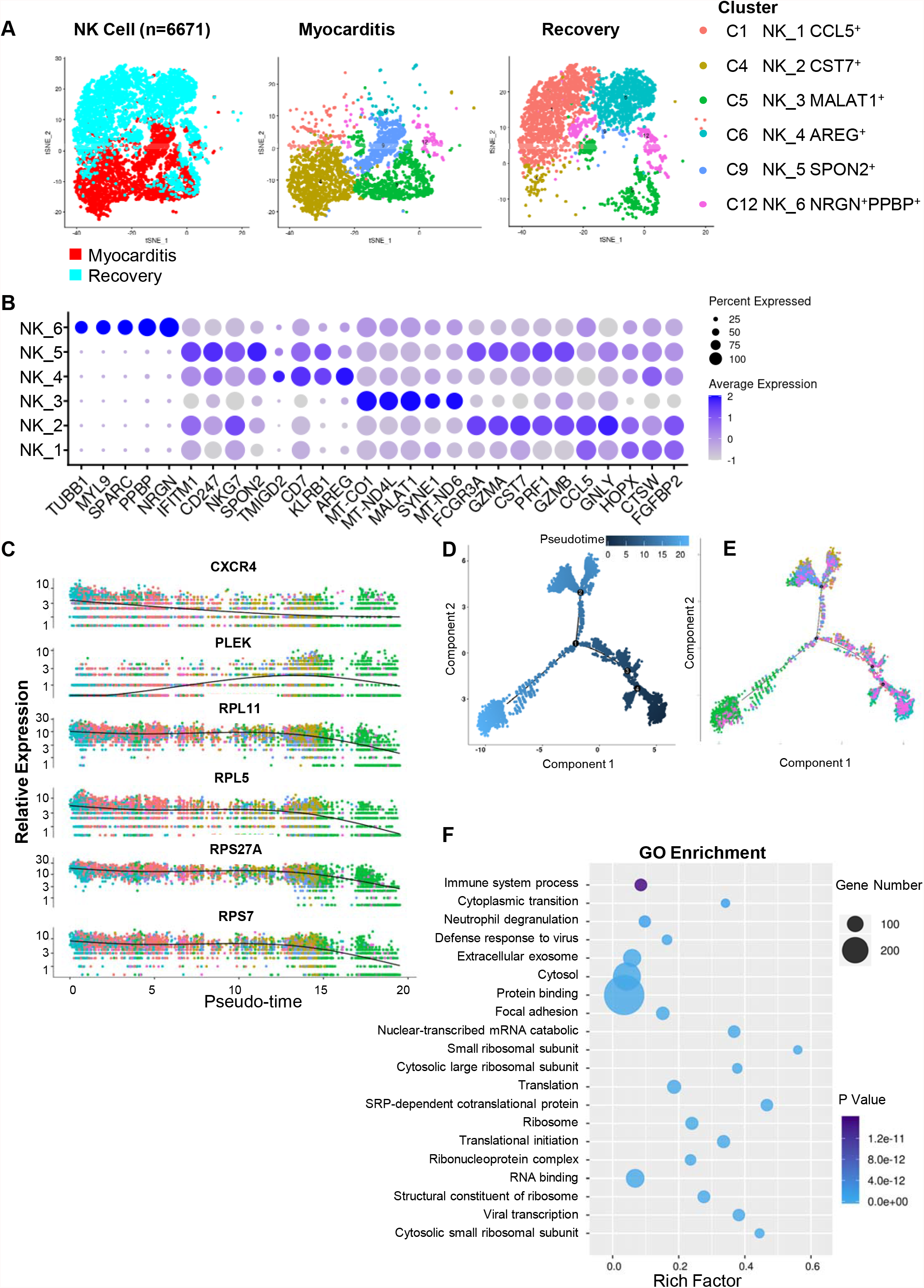
NK cell subpopulations and its function in ICI related myocarditis and recovery phase. A. tSNE plots of different NK cell subpopulations during the ICI myocarditis and recovery phase, each color corresponds to different NK cell subpopulation/subtype as the right panel showed; B. Dot plot showing the markers of each NK cell subpopulation. Dot size corresponds to proportion of cells within the group expressing each gene, and dot color corresponds to expression level; C.Trajectory analysis of NK cell using Mononcle2, dot color corresponds to pseudotime; D. Pseudotime trajectory analysis of six subtypes of NK cell by using Mono_2 (NEAT1^+^) as the root, each point indicates one single cell; E. Expressions of differential marker genes across six subtypes of NK cell; F. GO term enrichment analysis of main NK cell functions during ICI myocarditis.

Using trajectory analysis, we noticed that the C12 subpopulation (NK_6, NRGN^+^PPBP^+^) constituted a majority of the root of the trajectory, and C5 (NK_3 MALAT1^+^) from myocarditis located at the end of the trajectory (Figure 4, D). The gene expression distribution map with 6 predominately differential genes were shown in Figure 4, C; gradual decrease of CXCR4, RPL11, RPL5, RPS27A and RPS7, and increase of PLEK were identified with pseudotime. With the help of GO enrichment analysis, these NK cells mainly function in immune system processes, protein binding, RNA binding and neutrophil degranulation (Figure 4, D).

### T cell cluster may play an essential role in the PD-1 response

A total of 5072 T cells were identified. Unlike monocytes and NK cells, the subpopulation of T cells and its compositions remained generally the same during ICI related myocarditis and recovery phase (Figure 5, A). The mark genes of each subpopulations were shown in Figure 5, B. Previous report had identified four different types of gene mutations, which might be related to the efficacy of PD-1 antibody^12-19^. To further investigated the transcriptional changes of T cells in patients utilizing PD-1 inhibitors and suffering myocarditis, we then analyzed the expression of genes related to efficacy, resistance, and hyper-progressor in different T cell subpopulations. Notably, with regard to genes associated with PD-1 inhibitor resistance, JAK3, JAK1 and B2M were highly expressed in all T cell subpopulations, and PTEN in T cell_1 CD4^+^IL7R^+^, T cell _2 CD3D^+^CD3G^+^, T cell _3 CD27^+^AQP3^+^, and T cell _5 NRGN^+^PPBP^+^. The MDM4 related to hyper-progressor was also highly expressed in T cell_1 CD4^+^IL7R^+^ and T cell _3 CD27^+^AQP3^+^ both during myocarditis and recovery, and in T cell _5 NRGN^+^PPBP^+^ and T cell _4 MT-CYB^+^ during recovery (Figure 5, C).

**Figure 5.**
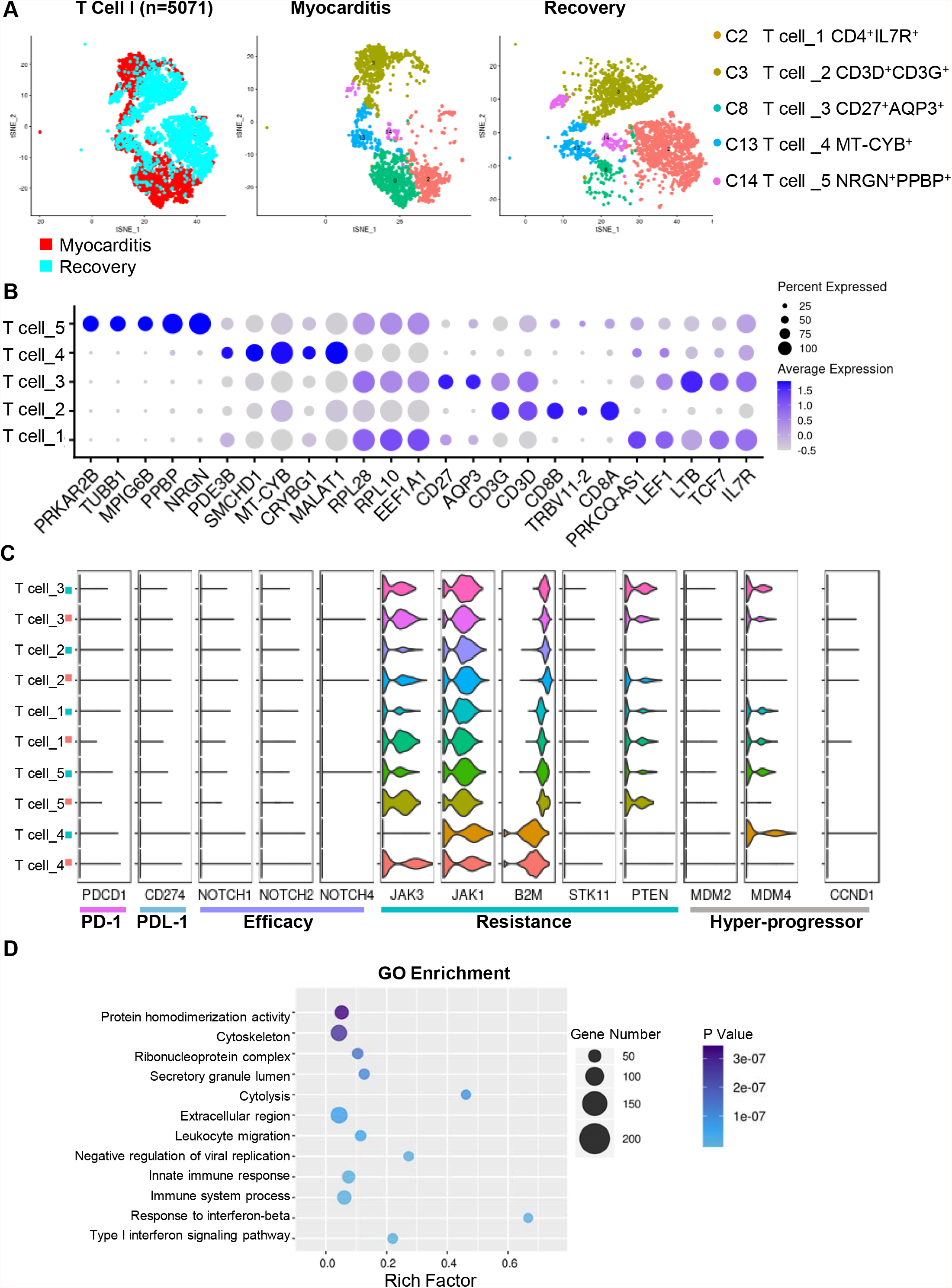
T cell subpopulations and the expression of genes related to PD-1 response. A. tSNE plots of different T cell subpopulations during the ICI myocarditis and recovery phase, each color corresponds to different T cell subpopulation/subtype as the right panel showed; B. Dot plot showing the markers of each T cell subpopulation. Dot size corresponds to proportion of cells within the group expressing each gene, and dot color corresponds to expression level; C.The expression of genes related to efficacy, resistance, and hyper-progressor in different T cell subpopulations; D. GO term enrichment analysis of main NK cell functions during ICI myocarditis.

### Substantial different BCR clone types were identified in ICI related myocarditis

B cells, although only 712 cells identified, were mostly prominently increased after recovery (Figure 6, A), suggesting that the humoral immunity was of vital importance for patients to recovery from ICI related myocarditis. As a result, we carried on to investigate the B cell receptor repertoire during myocarditis and recovery (Figure 6, B). The B cell receptors (BCRs) sequences of each single B cell were assembled from single-cell RNA-seq data using Cell ranger. Substantial different BCR clone types were identified with top 10 shown in Figure 6, C, suggesting that the BCR can react strongly during the ICI related myocarditis, along with B cell stimulation and clonal expansion. In the immune response, B cells could be activated by BCR recognition of antigen and lymphokines secreted by T cells to form effector B cells. There is a region called complementary determining region (CDR) on BCR, in which CDR3 is encoded by V, D and J genes, resulting in the highest diversity of CDR3. As a result, CDR3 V/J region genes could reflect the characteristics of BCR clonal typing. The abundance distribution of V/J gene (IGK, IGL, IGH) during myocarditis and recovery were shown in Figure 6, D. Substantial different V/J gene distributions were observed in ICI related myocarditis as compared to recovery phase.

**Figure 6.**
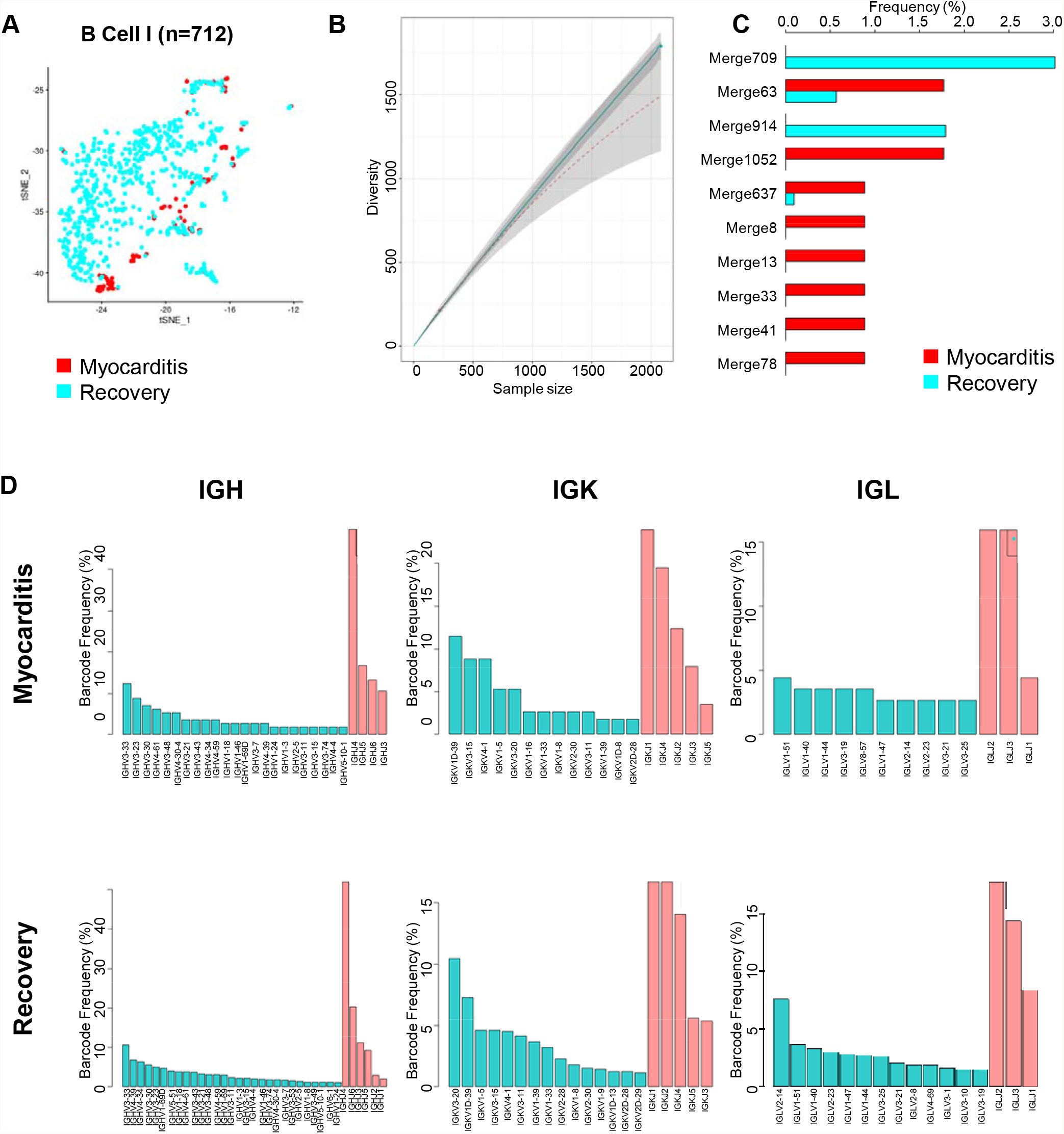
BCR clone types identified in ICI related myocarditis. A. tSNE plots of B cells during the ICI myocarditis and recovery phase; B. Diversity plot of B cell receptor repertoire during myocarditis and recovery; C. Top 10 substantial different BCR clone types were identified through BCRs sequences using Cell ranger; D. The abundance distribution of V/J gene (IGK, IGL, IGH) during myocarditis and recovery.

### Dysregulation of intercellular communication may appear in ICI related myocarditis

Intercellular communication is based on complex reactions between ligands and their receptors as well as specific cell signal pathways. By utilizing CellphoneDB, we built an intercellular network of potential ligand-receptor interactions among cell populations, aiming to analyze single-cell transcriptome data and explore cellular communication at a more detailed level. The overall intercellular communication between each pair of cell populations were shown in Figure 7, A and B. Six major cell populations (Monocytes, NK cells, CD4^+^ T cells, CD8^+^ T cells, B cells, and Megakaryocytes) have most communication with others in a complicated network. The top 20 protein interaction receptor-ligand pairs were shown in Figure 7, C. Of note, these intercellular communications, including TGFβ, HLA, CD74, etc. contributed to intercellular recognition and antigen presentation of the immune system. The disruption of intercellular communication may contribute to the pathogenesis as well as disease remission during ICI related myocarditis.

**Figure 7.**
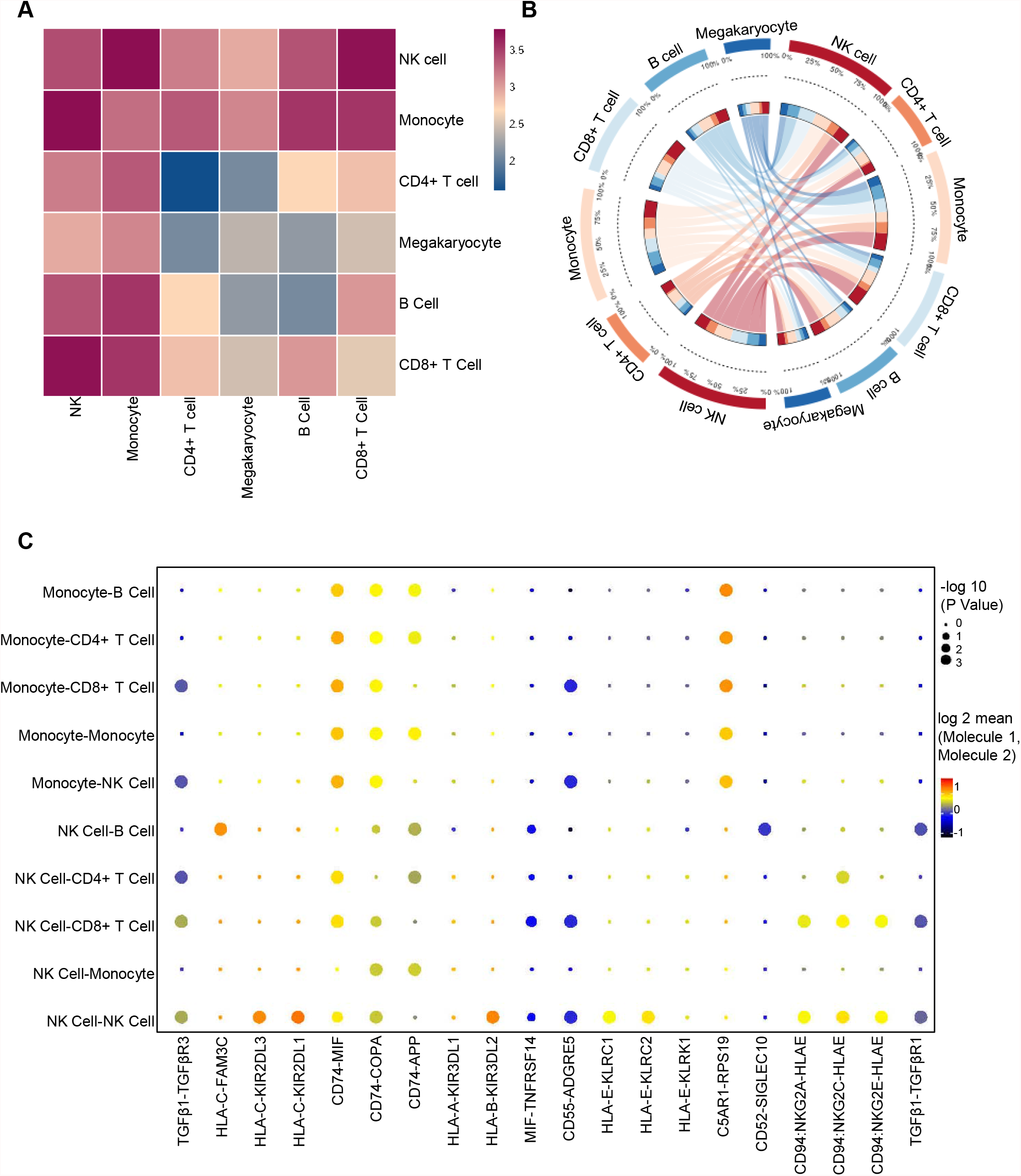
Dysregulation of intercellular communication in ICI related myocarditis. A. Overall intercellular communication between each pair of cell populations; B. Most ligand-receptor pairs communication among six major cell populations with others (Monocytes, NK cells, CD4^+^ T cells, CD8^+^ T cells, B cells, and Megakaryocytes) in a complicated network; C. The top20 protein interaction receptor-ligand pairs of different cell types between the ICI myocarditis and recovery phase.

## Discussion

Recently, multiple clinical studies have revealed potential tissue damage associated with ICI therapy; especially, treatment with ICIs can lead to severe and disabling inflammatory cardiovascular adverse-events soon after commencement of therapy^20^. However, few studies have been reported to obtain mechanistic insight into the molecular mechanisms of the ICI related myocarditis based on scRNA-seq of the immunes cells. In the present study, we have, to the best of our knowledge, for the first time presented the altered landscape of immune cells and differential genes in ICI related myocarditis during the disease activity and remission using scRNA-seq. Substantial immune cell composition and intercellular communication were found to be altered based on the following observations: (i) Monocyte, NK cell as well as B cell subpopulations contribute to the regulation of innate immunity and inflammation in ICI related myocarditis; (ii) T cell subpopulations highly express genes associated with PD-1 inhibitor resistance and hyper-progressor; (iii) Intercellular communication in ICI related myocarditis are significantly dysregulated.

ICIs have substantially improved clinical outcomes in multiple cancer types^21^. ICIs block proteins, such as CTLA-4, PD-1, or PD-L1 (PD-1 attached proteins) that help control the immune response. Blocking one of these proteins can release the “brake on the immune system” and enhance the ability of immune cells to attack tumor cells. However, blocking immune checkpoints to restore antitumor immune response may also break immune tolerance to self-antigens and induce specific immune-related adverse events (irAEs)^22^. The severity of most side effects is mild to moderate, which can be alleviated by general steroid treatment; but some patients may present with life-threatening side effects^23^. The irAEs highlight the fundamental difference between these drugs and other cancer treatments: conventional treatments such as chemotherapy directly kill tumor cells, while immunotherapy affect immune functions. As a result, irAEs can affect any organ and tissue. So far, our understanding towards irAEs is still limited, and it is still of great clinical challenge to predict the occurrence and severity of irAEs.

In the present study, we have provided a comprehensive spectrum and cell type– level resolution of peripheral blood cells during ICIs related myocarditis based on scRNA-seq. High throughput scRNA-seq can realize the isolation and library construction of tens of thousands of single cells, constructing a global tissue and cell transcription map. It is noteworthy that although PD-1 is mainly expressed in activated T cells and B cells, the subpopulations of monocyte and NK are more remarkably altered and exert their inflammatory function during ICI related myocarditis as compared to disease remission. These indicate that the tissue microenvironment affected by PD-1 inhibitor could cause dysregulated autoimmunity; and the loss of immune tolerance ultimately results in myocarditis.

ScRNA-seq has paved the way for the discovery of previously unknown cell types and subtypes in normal and diseased cardiovascular samples, facilitating the study of rare cells and the functional roles of immune cells in cardiovascular diseases such as myocarditis^24-26^. Although we are unable to describe each subtype and its biological functions in detail herein, it is worthy of attention that some key differential genes express in monocyte, NK cell and B cells instead of T cells. This indicated that the inflammatory monocyte played crucial role in the pathophysiological process of ICI related myocarditis. More importantly, the S100A protein family including S100A8, S100A9, S100A11 and S100A12 are significantly highly expressed in the monocyte subpopulation during ICI related myocarditis. Thus, it is also of clinical value to further investigate function of circulating S100A protein levels for predicting irAEs.

The increased B cell cluster together with substantially different B cell receptor repertoire suggest the innate immunity is of crucial importance during the acute disease onset, consistent with previous assumptions^1, 2^. The B cell receptor repertoire diversity is achieved by somatic recombination of immunoglobulin (Ig) genes centrally and by somatic hypermutation and Ig heavy chain class-switching peripherally^27^. Substantial differences during ICI related myocarditis indicate that there are altered functional gene rearrangements and gene combinations resulting in self-reactive BCRs as well as BCRs with high affinity for exogenous antigens after PD-1 inhibitor initiation. Hence, investigation of BCR repertoires based on larger population is reasonable to provide information on stages of B cell development and shed light on the etiology of B cell pathologies during irAEs.

The present scRNA-seq data have also revealed that the subpopulation of peripheral T cells and its compositions remained generally the same during ICI related myocarditis and recovery phase. However, the genes correlated to PD-1 inhibitor resistance and hyper-progressor have been highly expressed during myocarditis. This indicated that the resistance to PD-1 inhibitor treatment might be associated with high incidence of irAEs, and further clinical validation studies are necessary. On the other hand, previous study has also proposed that T-cell infiltration with CD4 and CD8 positivity is correlated to lethal myocarditis in combination with myositis in patients receiving nivolumab in combination with ipilimumab^28^. It is a pity that in the present study, we haven’t evaluated the single cell sequencing of the myocardium, and thus we have only researched the conclusion that the innate immunity has been extensively triggered during ICIs related myocarditis. But the single cell data from the myocardium of the patients exhibiting irAEs remain to be elucidated.

Intercellular communication analysis indicates that the intercellular recognition and antigen presentation differ during ICI related myocarditis and disease remission. Notably, transforming growth factor β (TGFβ) signaling pathways, involved in many cellular processes, including cell growth, cell differentiation and apoptosis take active part in the intercellular communication among NK cells, T cells and monocytes via binding TGFβR1 and TGFβR3. In addition, CD74, as an integral transmembrane molecule in intracellular sorting of MHC class II molecules also play an important role in intercellular communications. Generally, as genes associated with antigen presentation and autoimmune are important ligands, we reasoned that the disruption of intercellular communication in immune cells may contribute to the pathogenesis of irAEs. Previous studies have proved that targeting ligand-receptor interactions (for instance, with ICIs) can provide significant benefits for patients^29^. However, our knowledge regarding which interactions occur in a tumor and how these interactions affect outcome is still limited. Thus, our results shed light on possible downstream pathways and potential mechanism of irAEs which remain to be in-depth explored.

The main limitation of the present study is that it is a descriptive study. We have investigated the single cell profiling only in one patient previously using Tislelizumab due to the rare occurrence of the disease. However, by using the 10x Genomics platform to characterize the transcriptional profiles and functional pathways, we represent the first cell atlas during the onset of ICI related myocarditis and after disease remission, which could be used as a reference for ICI related studies in the future. More investigation should be done in the continuous exploration to reveal the roles of different immune cell populations for promoting and preventing the adverse events subsequent to immune checkpoint inhibitor utilization. By enrolling more samples and spatial information to scRNA-seq data, we believe that further in-depth investigation will improve our understanding of immune response and cell-to-cell interactions in ICI related adverse effects, so as to seek for novel diagnostic markers and targeted treatments.

In conclusion, by identifying altered pathways and highlighting a catalog of marker genes, this study has created a cell atlas of PBMC during ICI related myocarditis. We described the characteristics of cell populations, the respective marker genes and functions during the fulminant myocarditis and disease remission. In addition, we identified potential inflammatory cell populations and intercellular communication through vital ligand-receptor pairs, which would fuel advances in further mechanistic investigations. Gene expression changes in immune cell landscape provide a reference for organ toxicities related to ICI. Targeting specific immunological pathways represents a promising approach to treat ICI related myocarditis and other related toxicities. As the therapy of ICI related myocarditis is still limited in clinical practice, our investigation would shed light on the pathophysiological mechanism and potential therapeutic targets in continuous exploration.

### Ethics approval and consent to participate

Written informed consent was obtained from the patient, with ethnic committee approval at the First Affiliated Hospital of Xi’an Jiaotong University.

## Data Availability

All data produced in the present study are available upon reasonable request to the authors

## ABBREVIATIONS AND ACRONYMS

ICI: Immune checkpoint inhibitor
PBMC: Peripheral blood mononuclear cell
PD-1: Programmed Death Molecule 1
PD-L1: Programmed Cell Death 1 Ligand 1
TCR: T cell receptor
BCR: B cell receptor
CDR: Complementary determining region
irAEs: immune-related adverse events

## Acknowledgements

The authors wish to acknowledge the assistance of OmicStudio™ for the scRNA-seq and BCR repertoire analysis.

## Consent for publication

All authors have reviewed the final version of the manuscript and approve it for publication.

## Funding

This study was supported by National Natural Science Foundation of China (No. 82170464, 82100477), National Key R&D Program of China (2019YFA0802300, 2018YFC1311505), Key Research and Development Program of Shaanxi (No. 2021KWZ-25, S2020-YF-GHMS-0014, 2020GXLH-Y-015), Central University Basic Science Foundation of China (119132971000056); and the Clinical Research Award of the First Affiliated Hospital of Xi’an Jiaotong University, China (No. XJTU1AF-CRF-2018-025,XJTU1AF-CRF-2017-006).

## Authors’ contributions

JS, BZ and ZY participated in the design of the study. BL, MG FC and TZ collected the patients’ data and extracted the PBMC. BL, CW, GT and MG performed the statistical analysis. JS, BZ and BL drafted the manuscript. All authors approved the final manuscript.

## Conflicts of Interest

Not applicable.

## Figure legends

**Supplementary Figure 1.**
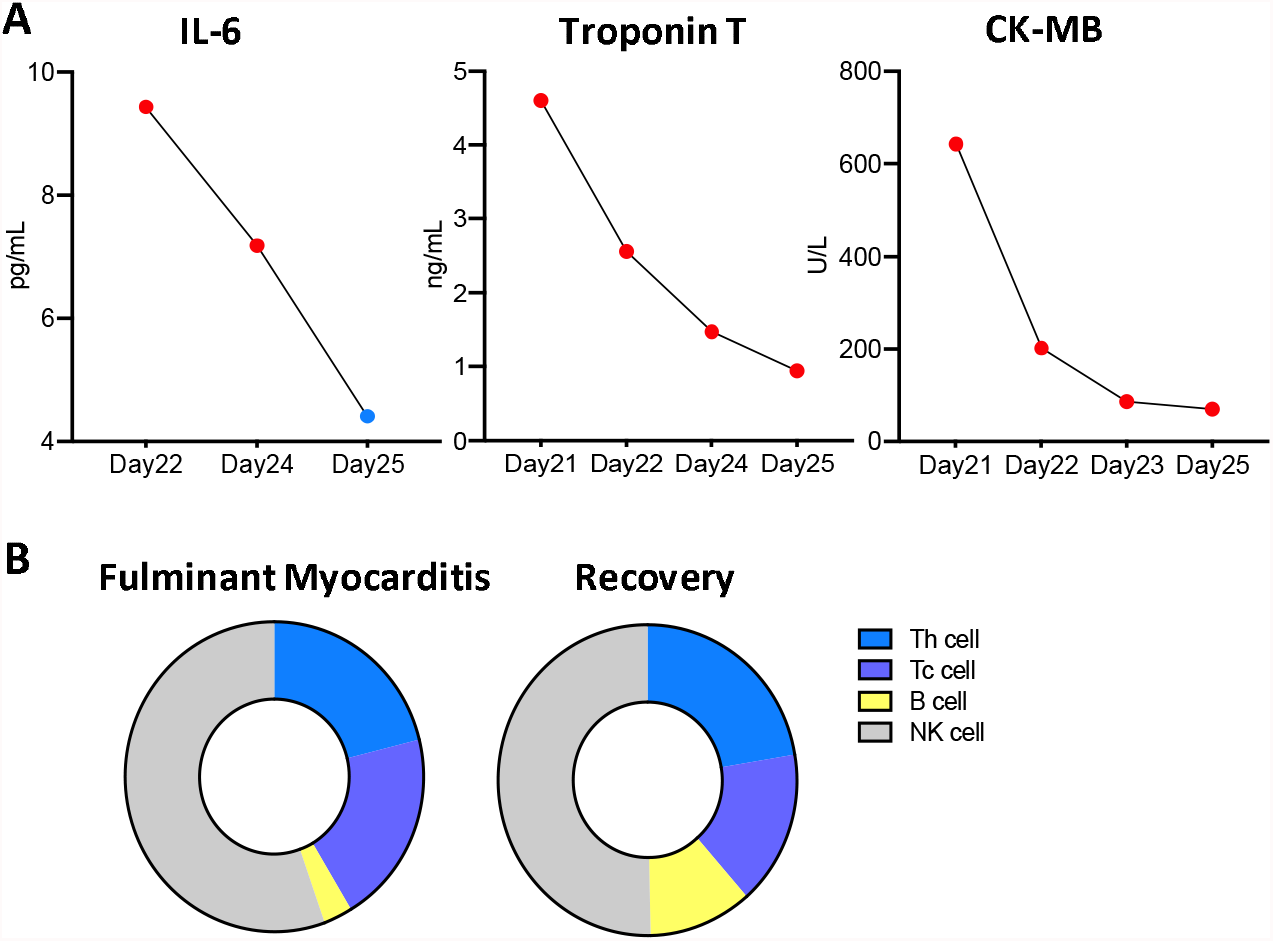
Clinical markers alteration during fulminant myocarditis and recovery. A. Levels of IL-6, Troponin T and CK-MB during the treatment; B. The immune subsets evaluated by the immunofluorescence assay.

**Supplementary Figure 2.**
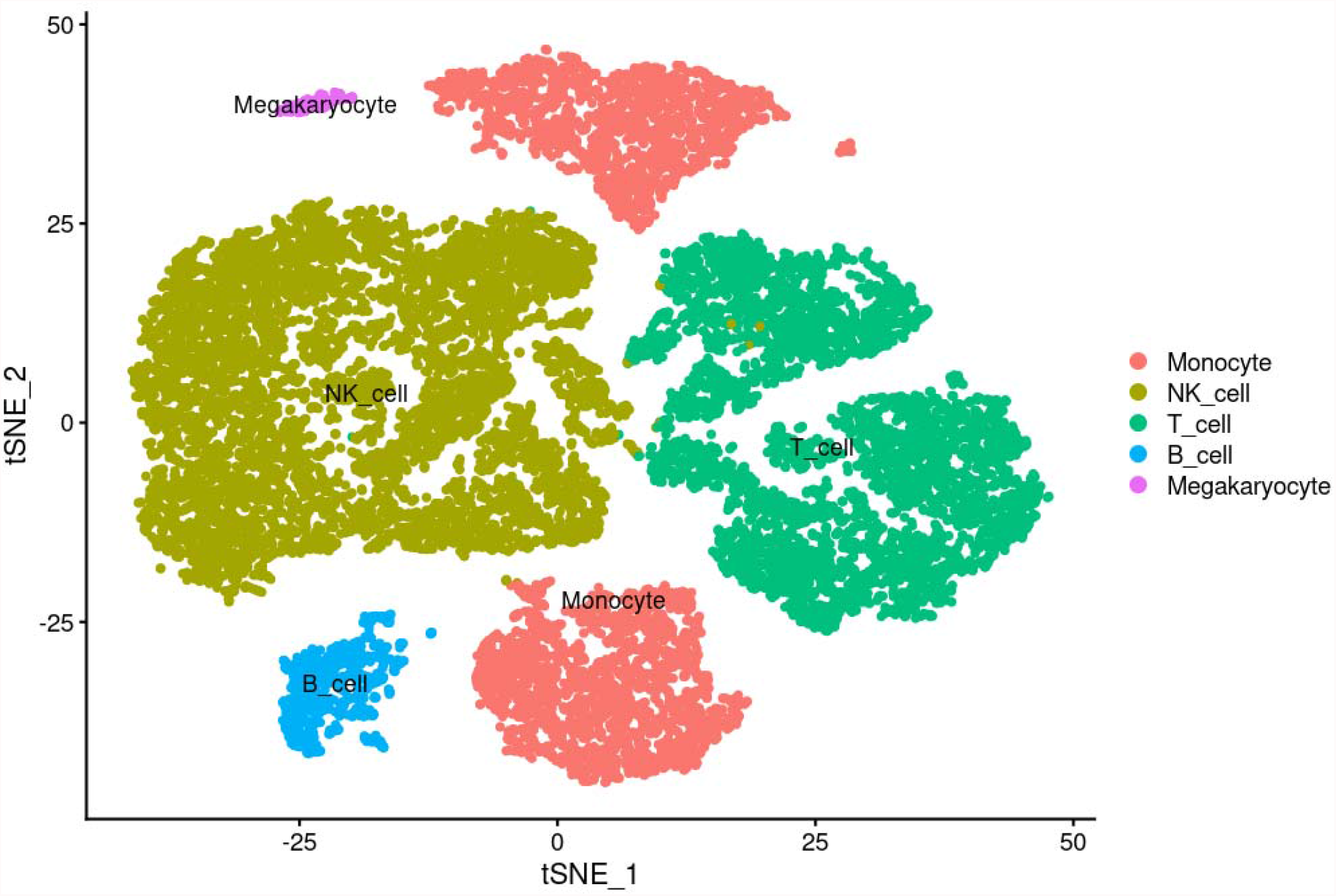
Clinical markers alteration during fulminant myocarditis and recovery. tSNE plot of clinical markers alteration during fulminant myocarditis and recovery using unsupervised graph-based clustering and SingleR.

## Notes

The authors declare no potential conflicts of interest.

### Competing Interest Statement

The authors have declared no competing interest.

### Author Declarations

Ethnic committee of the First Affiliated Hospital of Xian Jiaotong University gave ethical approval of this work

